# Longitudinal trajectories of depressive symptoms in children are influenced by baseline inflammation and HIV status

**DOI:** 10.1101/2025.11.12.25340107

**Authors:** Arish Mudra Rakshasa-Loots, Sarah K. Zalwango, Simon R. Cox, Alla Sikorskii, Bruno Giordani, Jorem E. Awadu, Amara E. Ezeamama

## Abstract

Mental health outcomes are substantially poorer among people with HIV than the general population. In a sample of 862 children and 439 adult caregivers in Kampala, Uganda, we investigated whether the trajectories of depressive symptoms over 24 months may be influenced by participants’ HIV status and baseline inflammation (indexed via high-sensitivity C-reactive protein, CRP). At higher baseline CRP concentrations, children with HIV showed lower baseline depressive symptoms relative to children without HIV. Over time, depressive symptoms increased in children with HIV but decreased in children without HIV. No differences in trajectories were observed in adults. Our results suggest that given high baseline inflammation, recovery from depressive symptoms may be significantly slower among children living with HIV compared to those without HIV. Specific interventions to reduce inflammation may need to be combined with more regular, holistic, and personalised interventions to alleviate depressive symptoms among children with HIV.

## Introduction

Inflammation is a notable predictor of mental health issues, including depression [1]. Increases in inflammatory biomarkers such as C-reactive protein (CRP) and interleukin-6 (IL-6) are associated with future depressive symptoms [2]. Inflammation has also been shown to affect the trajectories of depressive symptoms over time: for instance, one study of over 13,000 adults aged 50-90 years found that those with the highest levels of inflammation exhibited persistent and severe depressive symptoms across a 10-year follow-up [3]. In children and young people, as well, higher inflammation at baseline was recently shown to be associated with worse depressive symptom trajectories in two large, population-based cohorts [4]. Therefore, inflammation may significantly influence the course of depressive symptoms in both children and adults.

Nearly 40 million people are living with HIV globally [5]. Mental health outcomes are substantially poorer in this community than in the general population: up to two-thirds of people with HIV report mental health symptoms [6] and there is an approximately two-fold higher incidence for depression and other severe mental illnesses in this group [7]. Psychosocial and socioeconomic determinants such as HIV-related stigma, unemployment, and food or housing insecurity contribute significantly to the risk for depression in people with HIV [8]. However, it is also possible that biological factors such as inflammation may partly contribute to this risk [9], since people with HIV exhibit chronic low-grade inflammation even when receiving successful antiretroviral treatment (ART) [10]. Investigating the possible role of inflammation in depression amongst people with HIV thus serves a dual purpose: to identify potentially modifiable contributors to depression risk and deliver improved mental healthcare in this underserved community, as well as to develop a potential model for targeting ‘inflammatory depression’ in the wider population.

We have previously reviewed existing evidence on the associations of depression with biomarkers of inflammation in people with HIV [11]. In this scoping review, we identified certain critical gaps in the literature in this area: namely, most existing studies comprised primarily of older adults, men, and participants located in the US. This represents a major challenge to generalisability and translation of immunopsychiatry research for people with HIV, since the majority of people with HIV are located in eastern and southern Africa, and the median prevalence of HIV in these regions is higher among young women and girls [5].

In the current study, we sought to directly address some of these gaps. Using a large and gender-diverse sample of children and adults in Uganda, we aimed to determine whether the relationship between baseline inflammation and the longitudinal trajectories of depressive symptoms may be moderated by HIV status. We hypothesised that HIV status would significantly interact with baseline inflammation to influence depressive symptom trajectories.

## Methods

### Participants

This study involved a total of *N* = 1,301 participants, of whom *n* = 862 were children (291 without HIV, 292 perinatally exposed to but not living with HIV, 279 with perinatally acquired HIV) and *n* = 439 were adult caregivers (165 without HIV, 274 with HIV). Participants were recruited between June 2017 and December 2024 from the Kawaala Health Center IV (KHC-IV) in the Kawempe Division of Kampala, Uganda as part of three cohort studies in child-caregiver pairs. Inclusion criteria for children were: age 6-18 years at enrolment, having verifiable perinatal HIV status, and being accompanied by an adult (> 18 years old) caregiver with whom the child had resided for at least six months. For children or caregivers living with HIV, additional inclusion criteria were that they must be connected to HIV care at Kawaala Health Center (the study site) and the child-caregiver pair must reside within 25 kilometres of the Health Center. For children or caregivers not living with HIV, an additional inclusion criterion was providing consent to rapid diagnostic HIV testing to confirm their HIV status. Only children born in formal healthcare settings (e.g. a hospital) were enrolled, because their HIV and ART exposure status in pregnancy was objectively verifiable via medical records and the birth mother’s participation in perinatal HIV prevention programmes. All participants with HIV were receiving ART.

### Ethics approval

Adult participants provided written informed consent, and participants under the age of 18 provided written informed assent with written informed consent provided by their parent or caregiver. All study procedures were reviewed and approved by the Biomedical and Health Institute Review Board of Michigan State University (BIRB protocol references: 16-828, 205, and 5050), the Makerere University College of Health Sciences, School of Medicine for all studies at Kawaala Health Center (SOMREC protocol references: 2017-017, 2018-099 and 2021-85). In addition, the Uganda National Council for Science and Technology approved the protocol for all studies (protocol references: SS4378, HS 2466 and HS1532ES).

### Measures

Inflammation was measured at baseline using high-sensitivity C-reactive protein (CRP) concentrations in blood serum using standardized immunoassay methods, which reliably detect concentrations in the range relevant for both cardiovascular and neuropsychiatric research. Blood samples were centrifuged, and serum aliquots stored at *−*80°C until batch analysis. CRP was quantified using an enzyme-linked immunosorbent assay (ELISA) or particle-enhanced immunoturbidimetric assay, both of which are widely validated for epidemiologic studies. An empirical threshold of CRP greater than +1 SD above the mean (“high CRP”) and CRP lower than mean (“low CRP”) was used to categorise participants into two groups based on baseline CRP concentrations.

Depressive symptoms were assessed longitudinally, including at baseline and at six-monthly intervals for up to 24 months (maximum of five measurements per participant), using the nine-item Patient Health Questionnaire (PHQ-9). The PHQ-9 is a widely-used tool to assess depressive symptoms, and has been validated for use in Uganda, including with children and with people with HIV [12]. The PHQ-9 is scored between 0 and 27, with scores ≤ 4 indicating no or minimal depressive symptoms, and scores >10 typically showing acceptable sensitivity and specificity for detecting major depression [13].

### Statistical analysis

Statistical analysis was carried out using R (v4.4.1). Participant characteristics were summarised using medians and interquartile range (IQR) for continuous variables, and frequencies (n, %) for categorical variables. Comparisons of participant characteristics by HIV status were conducted using Wilcoxon’s rank-sum tests for continuous variables and Pearson’s chi-squared tests for categorical variables.

Latent growth curve modelling (LGCM) in a structural equation modelling (SEM) framework was carried out using R package `lavaan` (v.0.6-19) [14] to investigate trajectories of PHQ-9 scores, separately for children and adults. Given that SEM is sensitive to non-normality in data, PHQ-9 scores and CRP concentrations were log-transformed to reduce skewness (Supplementary Materials). Maximum likelihood estimation with robust standard errors (MLR estimator), which is robust to non-normality in data, was used in all models. Missing data were addressed using full-information maximum likelihood (FIML) estimation.

Unconditional LGCM was used to estimate the intercept (score at baseline) and slope (rate of change in score) for trajectories of PHQ-9 score. Model fit was assessed using the comparative fit index (CFI), Tucker–Lewis index (TLI), and root mean square error of approximation (RMSEA), with good fit indicated by values of CFI ≥ 0.95, TLI ≥ 0.95, and RMSEA ≤ 0.06 [15]. Conditional LGCM controlling for sex and age at baseline was then conducted to test for the main effects of HIV status and baseline CRP concentration, and the interaction between these predictors, on the intercept and slope of PHQ-9 scores as follows:

~~~
PHQ-9 score intercept ∼ CRP×HIV status + CRP
+ HIV status + age at baseline + sex
PHQ-9 score slope ∼ CRP×HIV status + CRP +
HIV status + age at baseline + sex
~~~

This model simultaneously gives estimates for the level and change in depressive symptoms, and the main and interaction effects of the two predictors (CRP and HIV status). The magnitude and significance of the interaction term was the primary outcome of interest. Main effects were determined from the full model (including the interaction term). Because there were three levels of HIV status among children, conditional LGCM models were also run separately for each pairwise combination of HIV status groups to identify which groups differed from each other (i.e. these were post-hoc tests to aid interpretation of the interaction results). Interactions between CRP concentrations and HIV status were visualised by categorising participants into high (CRP greater than +1 SD above the mean) and low (CRP lower than mean) baseline CRP groups. Standardised estimates with 95% confidence intervals (*β* [95% CI]) and *p* values were calculated for all models.

## Results

### Participant characteristics

Median (IQR) ages at baseline were 13 (11, 16) years among children and 39 (33, 47) years among adults. Overall, 53% of children and 89% of adults were female. At baseline, 91% of children and 80% of adults met criteria for low or no depressive symptoms (PHQ-9 score ≤ 4). Median (IQR) follow-up duration was 12 (0, 18) months among children and 6 (0, 18) months among adults, resulting in a total of *N* = 3,212 observations. Summary characteristics of participants stratified by HIV status are shown in **Table 1**. Sex distribution did not differ significantly between HIV groups, whereas participants with HIV were slightly older than other groups among both children and adults.

**Table 1.**
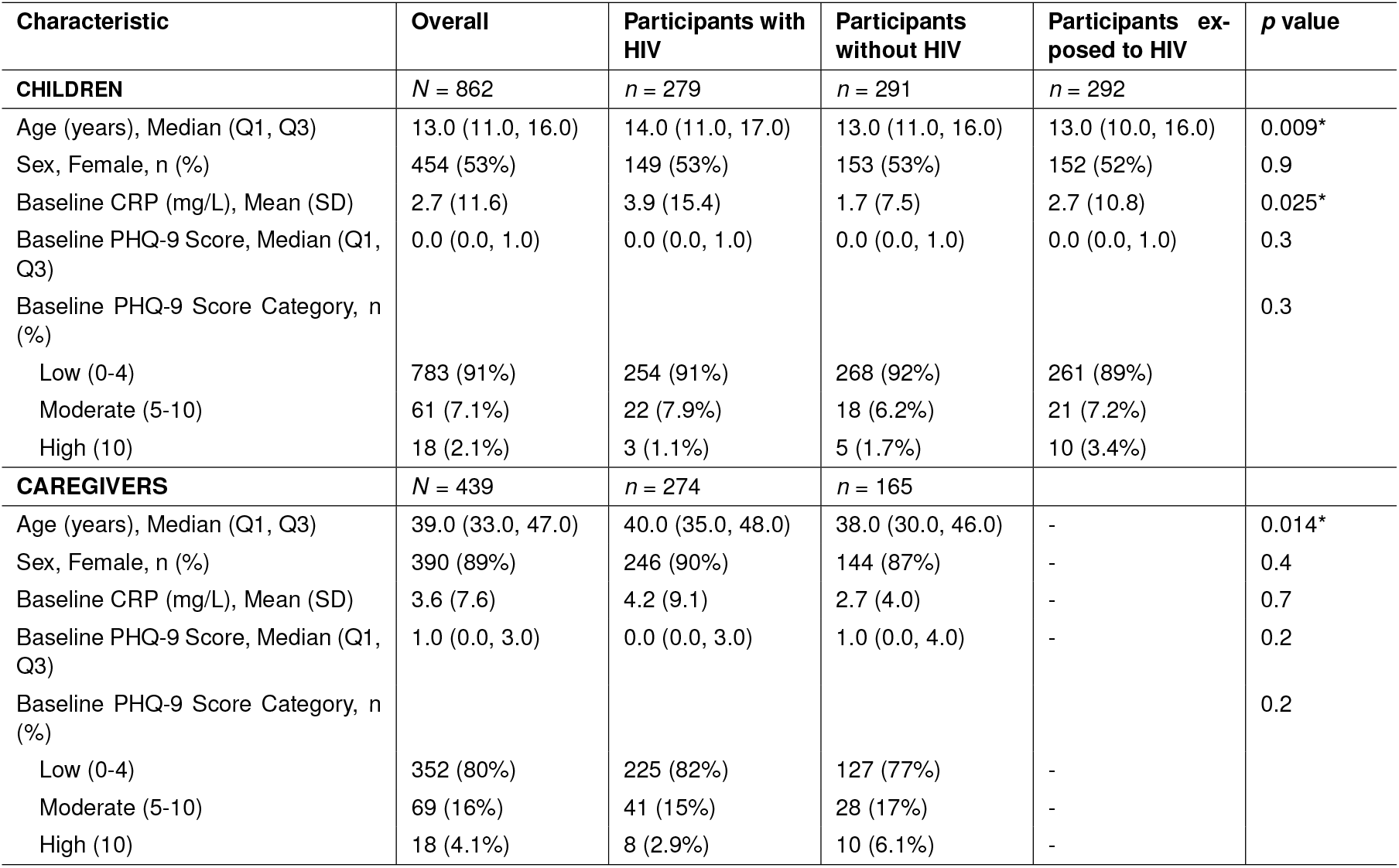
Participant characteristics. Comparisons between HIV status groups were conducted using Pearson’s chi-squared tests for categorical variables and Wilcoxon’s rank-sum tests for continuous variables. * indicates *p <* 0.05.

| Characteristic | Overall | Participants with HIV | Participants without HIV | Participants exposed to HIV | p value |
| --- | --- | --- | --- | --- | --- |
| **CHILDREN** | *N* = 862 | *n* = 279 | *n* = 291 | *n* = 292 |  |
| Age (years), Median (Q1, Q3) | 13.0 (11.0, 16.0) | 14.0 (11.0, 17.0) | 13.0 (11.0, 16.0) | 13.0 (10.0, 16.0) | 0.009* |
| Sex, Female, *n* (%) | 454 (53%) | 149 (53%) | 153 (53%) | 152 (52%) | 0.9 |
| Baseline CRP (mg/L), Mean (SD) | 2.7 (11.6) | 3.9 (15.4) | 1.7 (7.5) | 2.7 (10.8) | 0.025* |
| Baseline PHQ-9 Score, Median (Q1, Q3) | 0.0 (0.0, 1.0) | 0.0 (0.0, 1.0) | 0.0 (0.0, 1.0) | 0.0 (0.0, 1.0) | 0.3 |
| Baseline PHQ-9 Score Category, *n* (%) |  |  |  |  | 0.3 |
| Low (0-4) | 783 (91%) | 254 (91%) | 268 (92%) | 261 (89%) |  |
| Moderate (5-10) | 61 (7.1%) | 22 (7.9%) | 18 (6.2%) | 21 (7.2%) |  |
| High (10) | 18 (2.1%) | 3 (1.1%) | 5 (1.7%) | 10 (3.4%) |  |
| **CAREGIVERS** | *N* = 439 | *n* = 274 | *n* = 165 |  |  |
| Age (years), Median (Q1, Q3) | 39.0 (33.0, 47.0) | 40.0 (35.0, 48.0) | 38.0 (30.0, 46.0) | - | 0.014* |
| Sex, Female, *n* (%) | 390 (89%) | 246 (90%) | 144 (87%) | - | 0.4 |
| Baseline CRP (mg/L), Mean (SD) | 3.6 (7.6) | 4.2 (9.1) | 2.7 (4.0) | - | 0.7 |
| Baseline PHQ-9 Score, Median (Q1, Q3) | 1.0 (0.0, 3.0) | 0.0 (0.0, 3.0) | 1.0 (0.0, 4.0) | - | 0.2 |
| Baseline PHQ-9 Score Category, *n* (%) |  |  |  |  | 0.2 |
| Low (0-4) | 352 (80%) | 225 (82%) | 127 (77%) | - |  |
| Moderate (5-10) | 69 (16%) | 41 (15%) | 28 (17%) | - |  |
| High (10) | 18 (4.1%) | 8 (2.9%) | 10 (6.1%) | - |  |

### Trajectories of depressive symptoms

Overall, PHQ-9 scores at baseline were relatively low in both children (*β* = 0.18 [0.09, 0.27], *p <* 0.001) and adults (*β* = 0.25 [0.14, 0.35], *p <* 0.001), though slightly higher among adults. PHQ-9 scores decreased over time in both children (*β* = –0.17 [–0.23, –0.11], *p <* 0.001) and adults (*β* = –0.31 [–0.37, –0.24], *p <* 0.001), with a steeper decrease among adults (**Figure 1**). Model fit for a linear latent growth curve model was satisfactory for data from children (CFI = 0.91, TLI = 0.91, RMSEA = 0.06) and from adults (CFI = 0.95, TLI = 0.95, RMSEA = 0.06).

**Fig. 1.**
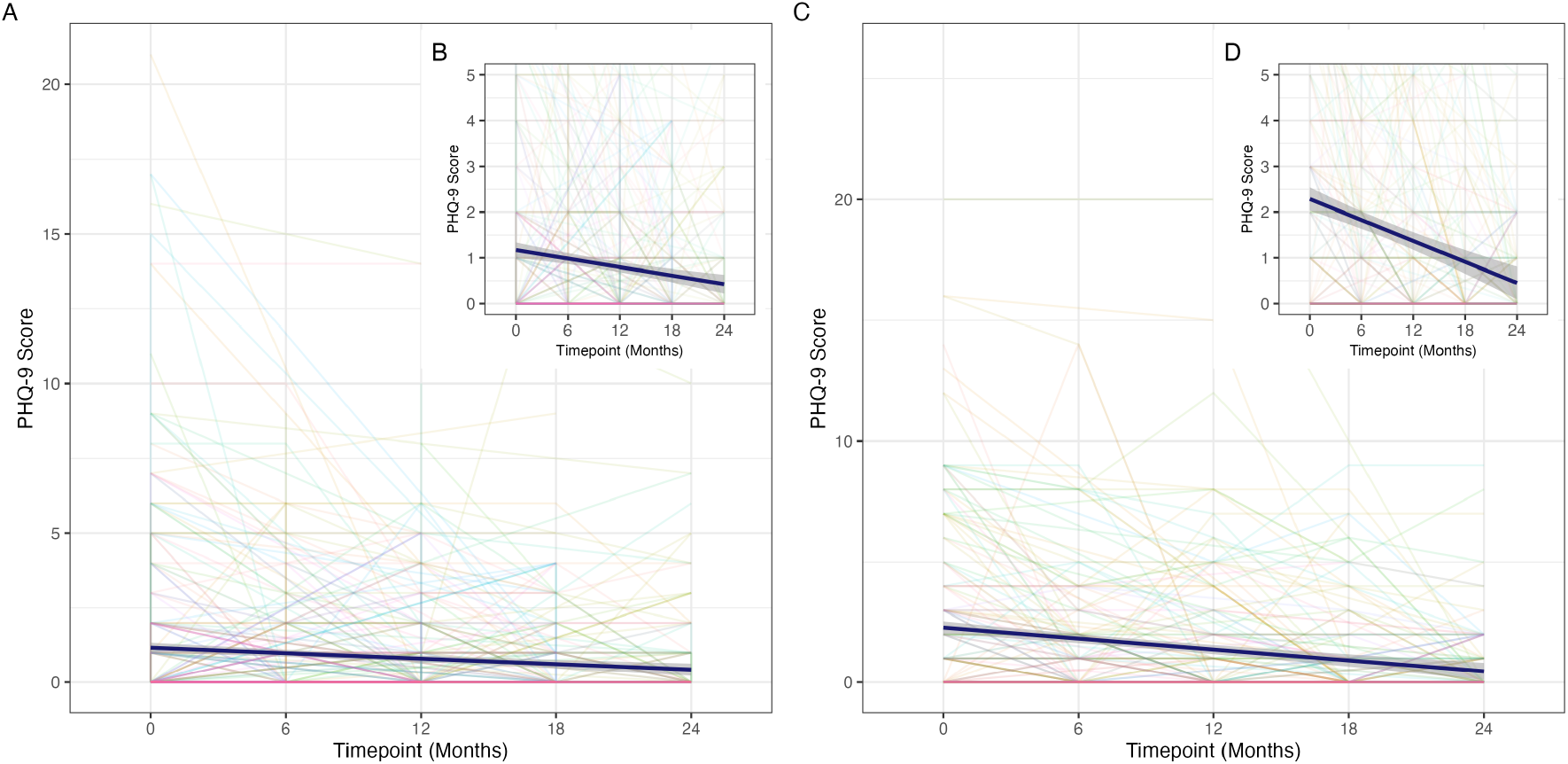
Trajectories of depressive symptom severity over time. Total scores on the Patient Health Questionnaire (PHQ-9) are shown for each participant (represented by a single line) over time, separately for **(A)** children and **(C)** adults. Mean trajectories with 95% confidence intervals for these groups are represented by dark blue lines. Given the relatively low severity of depressive symptoms in the sample overall, inset plots show a magnified view of the mean trajectories for **(B)** children and **(D)** adults to facilitate visual comparisons.

### Main effects of HIV status and baseline CRP

HIV status was not significantly associated with the intercept (*β* = 0.05 [–0.04, 0.14], *p* = 0.278) or slope (*β* = 0.03 [–0.03, 0.10], *p* = 0.293) of PHQ-9 score trajectories in children. Similarly, HIV status was not associated with the intercept (*β* = –0.08 [–0.18, 0.03], *p* = 0.159) or slope (*β* = 0.04 [–0.02, 0.11], *p* = 0.198) of PHQ-9 score trajectories in adults. Among children, baseline CRP concentrations also were not associated with the intercept (*β* = 0.07 [–0.02, 0.16], *p* = 0.143) of PHQ-9 score trajectories, but were nominally associated with the slope of these trajectories (*β* = –0.07 [–0.14, –0.001], *p* = 0.046). Among adults, baseline CRP was not significantly associated with the intercept (*β* = –0.05 [–0.15, 0.06], *p* = 0.367) or slope (*β* = 0.02 [–0.05, 0.08], *p* = 0.599) of PHQ-9 score trajectories.

### Interaction between HIV status and baseline CRP

Among children, the association of baseline CRP concentrations with the intercept (*β* = –0.09 [–0.18, –0.01], *p* = 0.034) and slope (*β* = 0.09 [0.03, 0.15], *p* = 0.004) of PHQ-9 score trajectories varied significantly according to perinatal HIV status. Pairwise comparisons revealed that this interaction was significant for children with HIV compared to children without HIV, as shown by the mean trajectories stratified by HIV status group plotted for participants with high and low baseline CRP concentrations in **Figure 2**. Among children with low baseline CRP, there was no difference in the intercept or slope of PHQ-9 scores based on perinatal HIV status. Among children with high baseline CRP, the intercept of PHQ-9 scores was lowest among children with perinatally acquired HIV and highest among children without HIV (*β* = –0.13 [–0.23, –0.02], *p* = 0.019). Children without HIV and children exposed to HIV exhibited a steady decline in PHQ-9 scores over time. However, children with HIV showed an overall increase in PHQ-9 scores over time (*β* = 0.11 [0.03, 0.19], *p* = 0.006) relative to children without HIV. Among adults, HIV status did not significant influence the association of baseline CRP concentrations with either the intercept (*β* = 0.07 [–0.04, 0.18], *p* = 0.224) or the slope (*β* = 0.04 [–0.03, 0.11], *p* = 0.260) of PHQ-9 score trajectories.

**Fig. 2.**
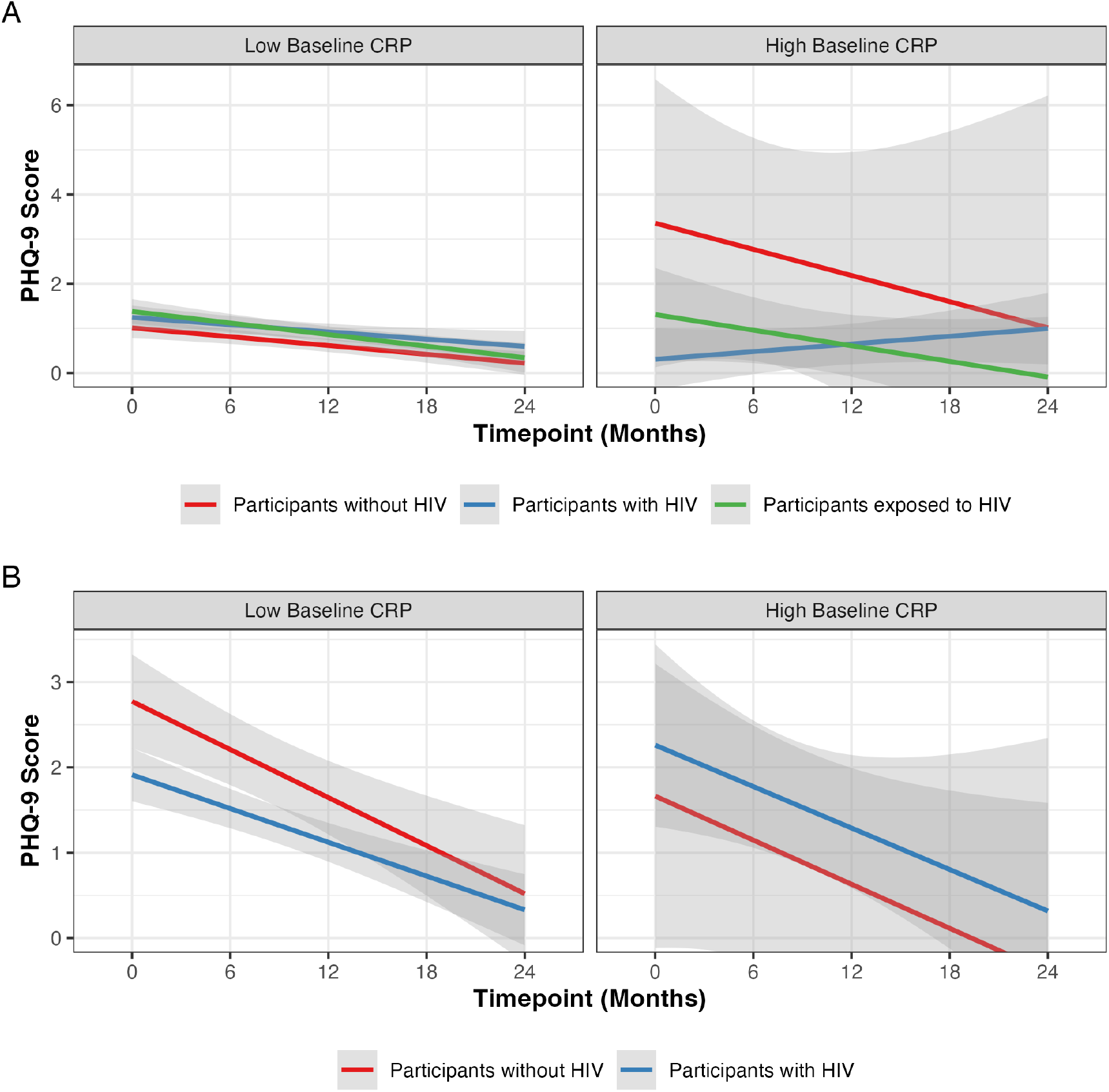
Mean longitudinal trajectories of depressive symptoms as functions of HIV status and baseline CRP concentrations among (A) children and (B) adults. Participants with baseline CRP concentrations greater than +1 SD above the group mean were categorised as having “High Baseline CRP” and those with CRP concentrations lower than the group mean were categorised as having “Low Baseline CRP”. Individual lines represent the fitted linear models with shaded grey areas representing 95% confidence intervals.

## Discussion

In this study involving 1,301 participants followed for up to 24 months in Uganda, we found that the relationship between inflammation and trajectories of depressive symptoms varied significantly based on HIV status in children, but not in adults. Children with HIV who exhibited higher baseline CRP concentration had lower starting PHQ-9 scores, and their scores increased significantly over time, relative to the PHQ-9 score trajectories of children without HIV (whose PHQ-9 scores started higher but tended to decrease over time). Therefore, at higher CRP concentrations, children with HIV in this study showed lower baseline depressive symptoms compared to children without HIV, but their symptoms increased over time.

Previous research in the general population has revealed small but significant associations between inflammation (as measured by CRP) and future depression in children[16]. In line with previous studies involving young people with HIV, we observed that depressive symptoms in participants generally decreased over time [17]. To our knowledge, this is the first study to investigate whether the trajectory of depressive symptoms over time in people with and without HIV differ in relation to baseline inflammation. In this sample, we observed that at higher baseline CRP concentrations, children with HIV on average reported lower baseline depressive symptoms than children without HIV. Given that children with perinatally acquired HIV have lived lifelong with chronic HIV morbidity, what at first appears as a counter-intuitive finding may reflect relatively higher emotional adjustment in this population navigating life with chronic morbidity that is absent in children without HIV. Higher self-reported depressive symptoms at baseline among children without HIV compared to those living with HIV (at comparable physiologic dysregulation) aligns with well-described response-shift phenomena in chronic illness, whereby internal standards and the meaning of symptom ratings change with adaptation to persistent morbidity [18].

This adaptation contributes to the disability paradox (good self-reported quality of life despite severe illness) [19] and to reporting heterogeneity across groups with different health expectations [20]. Similar valuation gaps are seen when patients and healthy respondents appraise identical health states, suggesting that appraisal as well as biology may drive self-reported baseline depressive samples in this study.

Crucially, we found that the main effect of HIV status on the change in depressive symptoms was not significant (consistent with previous studies) [21], but depressive symptoms tended to increase over time in children with HIV who had higher baseline CRP concentrations. Therefore, a key conclusion from the current study is that when considering the impact of HIV status on change in mental health outcomes such as depression, careful attention must be paid to potential for fundamentally different reference for physical and mental wellbeing in people with and without HIV. In other words, HIV status may be a key indicator that drives the trajectory of mental health outcomes, with change over time best understood when accounting for other risk factors such as inflammation.

Some important limitations of the study are noted. This study did not involve recruitment from psychiatric clinics; as a result, the severity of depressive symptoms in this sample was low overall, with only a small proportion of participants meeting criteria for severe depressive symptoms. Relatedly, depressive symptoms were measured using a self-reported questionnaire, which – although well-validated to screen for depressive symptoms – does not provide a gold-standard clinical assessment. CRP concentrations were only measured at baseline, and thus we could not investigate the association of interest in the reverse direction, i.e. between baseline depressive symptom severity and longitudinal changes in inflammation. Additionally, CRP concentrations in blood represent a highly dynamic measure of acute inflammation, and could be influenced by a wide range of factors such as a recent infectious exposure, body mass index, or alcohol use [22]. Non-normality in the distribution of PHQ-9 scores and baseline CRP may also have impacted the growth models, although we sought to account for this using log-transformation of these variables and employing robust estimators in LGCM.

Despite these limitations, this study also benefits from several strengths which provide a novel and important contribution to the literature. The sample size of this study is substantially larger than most previous studies in the field [11], resulting in sufficient statistical power to model complex relationships with high precision. Previous studies have also been largely cross-sectional, whereas this study involved repeated measurement of depressive symptoms, enabling the modelling of change in depressive symptoms over time. Finally, this study directly addresses critical gaps in the field by using data from a gender-diverse sample of individuals in an African setting. Given that the majority of people living with HIV are women and located in eastern and southern Africa, it is essential that research on mental health outcomes in people with HIV is carried out in these populations to maximise generalisability and impact.

Future research may seek to extend our findings by determining whether other biomarkers of inflammation, such as the cytokines TNF-α and IL-6, may similarly interact with HIV status to influence the trajectories of depressive symptoms. Epigenetic (DNA methylation) signatures of CRP represent a more longitudinally stable measure of inflammation than protein quantification in blood [23]. These epigenetic signatures may also be promising biomarkers to better understand whether chronic inflammation (rather than acute inflammation) influences the trajectories of depressive symptoms in children with HIV. Finally, it will be useful to explore these relationships in clinical (psychiatric) samples, including people with severe depression, bipolar disorder, or psychosis, in addition to community-based samples.

In summary, our results suggest that depressive symptoms may be less pronounced initially – but increase over time – in children with HIV, compared to children without HIV, for those with high baseline inflammation. These findings have important potential implications for clinical management and intervention for depressive symptoms among children with HIV. Our results suggest that given high systemic inflammation, recovery from depressive symptoms may be significantly slower among children living with HIV compared to those without HIV. Therefore, specific interventions to reduce inflammation may need to be combined with more regular, holistic, and personalised interventions to alleviate depressive symptoms among children with HIV. Future experimental medicine studies should also seek to determine whether reducing inflammation through targeted immunotherapies may partly contribute to a reduction in depressive symptoms in this population.

## AUTHOR CONTRIBUTIONS

AMRL analysed data and performed statistical analysis with input from SRC and AS; and AMRL wrote the first draft of the paper. AEE designed research; AEE, SKZ, BG conducted research. All authors edited the manuscript for critically important content and approved final version of the manuscript.

## ACKNOWLEDGEMENTS

We thank all the research participants who contributed their time and data to this study. We acknowledge the dedication and hard work of the field research staff which enabled data collection: Dr Lois Bayigga, Ms Gloria Nakigudde, Ms Esther Nakayenga, Mr Arnold Katta, Ms Ruth Nalwoga, Ms Phiona Nalubowa, Ms Faridah Nakatya, and Ms Irene Asiingura. AMRL thanks Prof Barbara Laughton for input into earlier versions of this study.

## DATA AVAILABILITY

Data described in the manuscript, code book, and analytic code will be made available upon request pending application clarifying intended use and subject to a formal data use agreement. Analysis code developed for this study is available publicly at github.com/arishmr/depr-traj-uganda.

## COPYRIGHT STATEMENT

For the purpose of open access, the authors have applied a CC-BY public copyright licence to any Author Accepted Manuscript version arising from this submission.

